# Resting-state electroencephalography microstates in antipsychotic-naïve individuals across the psychosis spectrum

**DOI:** 10.64898/2025.11.28.25341198

**Authors:** Fabian M. Mager, Tina D. Kristensen, Edwin van Dellen, Livia S. Dominicus, Mette Ø. Nielsen, Kirsten B. Bojesen, Cecilie K. Lemvigh, Mikkel E. Sørensen, Dorte Nordholm, Birgitte Fagerlund, Merete Nordentoft, Louise B. Glenthøj, Birte Y. Glenthøj, Bob Oranje, Lars K. Hansen, Bjørn H. Ebdrup, Karen S. Ambrosen

## Abstract

Aberrant microstate features of resting-state electroencephalography (rsEEG) have been proposed as potential endophenotypic markers for schizophrenia. However, longitudinal investigations across the psychosis continuum remain limited, particularly regarding treatment effects and illness-related alterations over time.

We examined 47 antipsychotic-naïve, first-episode patients with psychosis (FEP), 32 individuals at ultra-high risk for psychosis (UHR), and 94 matched healthy controls (HC). All participants underwent rsEEG at baseline, FEP patients and HC were reassessed after six weeks and two years, during which the FEP-patients received antipsychotic treatment. We analyzed microstate features: Duration, Occurrence, Coverage, and Sample Entropy, and investigated group differences at baseline, as well as within group and between group effects over time. Additionally, we explored the effect of medication, and associations with symptom level using a linear model. Post hoc correlation analyses were performed to investigate the stability of microstate features at an individual level over time.

At baseline, no differences were observed among HC, UHR, and FEP groups. The main linear mixed models including HC and FEP, as well as all microstates A, B, C, and D at the 3 timepoints indicated an overall effect of time between baseline and two years for Occurrence (*p*=0.044), and Coverage (*p*=0.036), but not for Duration (*p*=0.986). Post-hoc tests for each microstate showed a significant effect of time within FEP between baseline and two years (Occurrence *p*=0.047, Duration C *p*=0.011, and Coverage C *p*=0.007). Furthermore, a group effect emerged between HC and FEP at two years for Coverage C (*p*=0.039), p-values uncorrected. No other effects of time or group were observed. Occurrence of Microstate D was negatively correlated with general symptom level at baseline (*p*=0.008, corrected), but not at any follow-up. No associations were found between microstates and antipsychotic medication at six weeks. These findings indicate that Microstate C increases in antipsychotic naïve patients the first two years after first episode psychosis, contrasted with the temporal stability in controls. This highlights the need for further research disentangling longitudinal effects of pharmacological and pathophysiological modulation of EEG microstates in large samples.

## Background

Schizophrenia is a severe mental disorder, and early detection is vital as it may facilitate timely interventions mitigating illness progression, influencing disease trajectory and improvement of long-term outcomes^1,2^. Identification of reliable biomarkers present in the very early stage of the disease could be used to guide treatment, potentially improving the outcome by protecting against neurobiological changes associated with the illness^3^.

Neurophysiological aberrations assessed with electroencephalography (EEG) in patients in the chronic phase of illness are well-established^4^. Aberrations of EEG microstates have been proposed as an endophenotype for schizophrenia^5^, potentially useful for early detection and treatment monitoring^6^. EEG microstates are derived from resting-state brain electrical activity and constitute discrete, recurrent scalp potential configurations^7^, which can reliably be used as neurophysiological features^8^. Four microstate topographies are consistently identified across healthy individuals and are commonly labeled microstate class *A, B, C,* and *D*^9,10^. Three temporal features can be derived for each microstate class: Duration, Occurrence, and Coverage. Duration is the average time a given microstate persists before transitioning; Occurrence is the average number of occurrences of a given microstate per second; and Coverage in percent is the time spent in a microstate divided by the total recording time.

Previous studies have suggested that increased presence of microstate *C* and decreased presence of microstate *D* may characterize patients with schizophrenia^11^ as well as their unaffected siblings^5^. However, this current evidence is commonly based on small samples of medicated and predominantly chronically ill patients, leaving the question of how antipsychotic medication and chronicity may affect microstates *C* and *D* unanswered.

Besides the three temporal features (Duration, Occurrence, and Coverage), microstates can be characterized by their sequence information, i.e., the transitional statistics from one microstate to another. Microstate Sample Entropy is a measure of sequence predictability^12^ which has recently been reported to show less predictable microstate sequences in medicated early-course psychosis patients compared to healthy controls (HC)^13^, where the degree of randomness decreased with increasing medication dose. To our knowledge, no studies have investigated microstate sample entropy in antipsychotic-naïve patients in the psychosis spectrum.

Biomarkers present even before illness onset may provide insight into underlying neurobiological mechanisms contributing to schizophrenia. However, biomarkers cannot be understood in isolation and need to be associated with clinical features, such as core symptoms. In the present study, we investigated microstate temporal parameters at baseline in antipsychotic-naïve patients across the psychosis continuum, comparing HC, individuals at ultra-high risk of psychosis (UHR), and patients with first-episode psychosis (FEP). Based on the studies mentioned above^5,11^, we hypothesized increased microstate C and a decreased microstate D in FEP patients compared to UHR and HC. Our secondary aim was to examine the within- and between group effect of microstate temporal parameters over time, from baseline to 6 weeks and 2 years follow up, as well as from 6 weeks to 2 years follow up in HC and FEP. Finally, we explored if potential changes over time could be explained by medication and/or illness progression.

## Methods

### Participants

FEP and UHR patients were recruited from in- and outpatient clinics in the Capital Region of Copenhagen, Denmark. FEP and HC were collected as part of two consecutive cohorts: Pan European Collaboration on Antipsychotic-Naïve Schizophrenia, PECANS (recruited from in- and outpatient facilities from December 17^th^ 2008 until January 14^th^ 2016), ClinicalTrials.gov Identifier: NCT01154829, and described in Nielsen et al^14^; and the PECANSII, (recruited from in- and outpatient facilities from January 2nd 2014 until January 28^th^ 2020), ClinicalTrials.gov Identifier: NCT02339844, described in Bojesen et al^15,16^. The studies were approved by the Regional Danish Committee on Health Research Ethics (H-D-2008-088, H-3-2013-149). FEP data were collected between 2008 and 2020. UHR patients were recruited by the Mental Health Center Copenhagen from in- and outpatient facilities from 2009 until 2013, approved by the Regional Ethics Committee of the Capital Region, Denmark (H-D-2009-013)^17^ and described in Madsen et al^18^. All participants provided written informed consent.

In this study, we included baseline data from antipsychotic-naïve first-episode patients with schizophrenia or other primary psychotic disorders (FEP_0_=47), patients with ultra-high-risk for psychosis (UHR_0_=32), and healthy controls (HC_0_=94). FEP and HC were followed up at 6 weeks (trial cessation) and 2 years after baseline (HC_6_=85, HC_2Y_=47, FEP_6_=24, FEP_2Y_=19). First-episode patients were part of clinical trials, receiving monotherapy (tool compound) for 6 weeks, either amisulpride (PECANS) or aripiprazole (PECANSII).

### Clinical Assessments

In the PECANS studies, ICD-10 diagnoses of schizophrenia were ascertained by Schedules for Clinical Assessment in Neuropsychiatry (SCAN 2.0). Patients with a diagnosis of schizophrenia or other primary psychotic disorders were included in this study (43 subjects with F20x and 4 subjects with F2x). UHR patients fulfilled one or more criteria of the Comprehensive Assessment of At-Risk Mental States (CAARMS)^19^: sufficient intensity and frequency of attenuated psychotic symptoms, and/or brief limited intermittent psychotic symptoms, and/or the vulnerability criteria (a first-degree relative with psychotic disorder or a diagnosis of schizotypal personality disorder), along with either a sustained low function (≤50) for at least one year or declining functioning (≤30%) for at least one month, as measured by the Social and Occupational Assessment Scale (SOFAS)^20^. The level of functioning of FEP was measured using the Global Assessment of Functioning Scale (GAF), the Social and Occupational Assessment Scale (SOFAS) [20]. The Positive And Negative Syndrome Scale (PANSS)^21^ were used to measure the level of psychotic symptoms of FEP (PANSS Total score, Positive, Negative, General). Medication was assessed by calculating the chlorpromazine equivalent dose (CPZ_Eq) for of each prescribed compound at week 6 and after 2 years.

### EEG acquisition

As part of the large multimodal studies, the participants were examined with the Copenhagen Psychophysiology Test Battery^22,23^. EEG was recorded using 64 *ActiveTwo* electrodes (BioSemi B. V., Amsterdam, The Netherlands) placed according to the extended 10-20 system with a sampling frequency of 2048 Hz. During acquisition, all participants were seated in a sound-insulated room (40 dB). After completing event-related paradigms, participants underwent 10 minutes of continuous resting-state EEG. During the resting-state acquisition, participants were instructed to sit still, relax, and keep their eyes closed. We requested participants to abstain from caffeine on the test day and nicotine intake one hour prior to the EEG assessments. Urine was screened for cannabis, cocaine, opiates, and amphetamines on the day of testing (Syva® RapidTest d.a.u® 4). Microstates analyses were based on the resting state recordings.

### Preprocessing of EEG recordings

Preprocessing and microstate analysis of resting-state EEG was performed in MATLAB® (2018b, The MathWorks Inc., Natick, Massachusetts, USA) using EEGlab^24^ and an open-source microstate plugin^25^. From the original 10-minute recording, we disregarded the first 30 and the last 90 seconds, resulting in 8 minutes of data per individual. Data were down-sampled and bandpass-filtered to 1-40 Hz. Noisy and largely artifacted channels and poor-quality portions of data were removed using EEGlab’s build-in automatic artifact rejection, and data were partitioned into continuous 4-second epochs. In addition, we used Independent Component (IC) analysis and an IC-classifier^26^ to remove eye-, muscle-, movement-, heart-, and noise artifacts. After the removal of artifacts, missing channels were interpolated. The code for preprocessing and analysis, including hyperparameters for artifact removal, is available from https://github.com/fmager/Microstates. The average number of epochs across all participants was 93.5, corresponding to 6.2 minutes of recording. The number of epochs and the number of interpolated channels did not differ between groups (*p*=0.58 and *p*=0.82, respectively).

### Microstate analyses

An overview of the pipeline is shown in Figure 1. For each subject, we entered scalp maps at random Global-Field-Power (GFP) peaks into a modified K-means algorithm^27^. GFP is a measure of global brain activity, and high GFP indicates a high signal-to-noise ratio (SNR)^28^. The procedure was repeated 6 times to retrieve sets of 4 to 10 microstates (*K* = {4, 5, …, 10}). Each set of microstates was compared to four template microstates from de Cruz et al.^5^, using spatial correlation (ρ ) as a similarity measure calculated with Global Map Dissimilarity (GMD)^28^. We matched microstates using a ‘winner-takes-it-all’ strategy and labeled them accordingly from *A* to *D*. When the number of microstates exceeded the number of template microstates (*K*>4), the remaining microstates remained randomly ordered. The polarity of microstate scalp maps during matching was ignored. After calculating spatial correlation for all microstates in each set and identifying microstates *A*, *B*, *C,* and *D*, we estimated the optimal number of microstates by comparing the first four microstates *A* to *D* in each set of microstates with the template microstates. To ensure similar microstate scalp topographies across groups, we performed a repeated-measure ANOVA (rmANOVA) of spatial correlations of microstates *A* to *D* with their respective template microstates using microstate class as within-subject and group as between-subject factor (Supplementary Text S1 for details).

**Figure 1:**
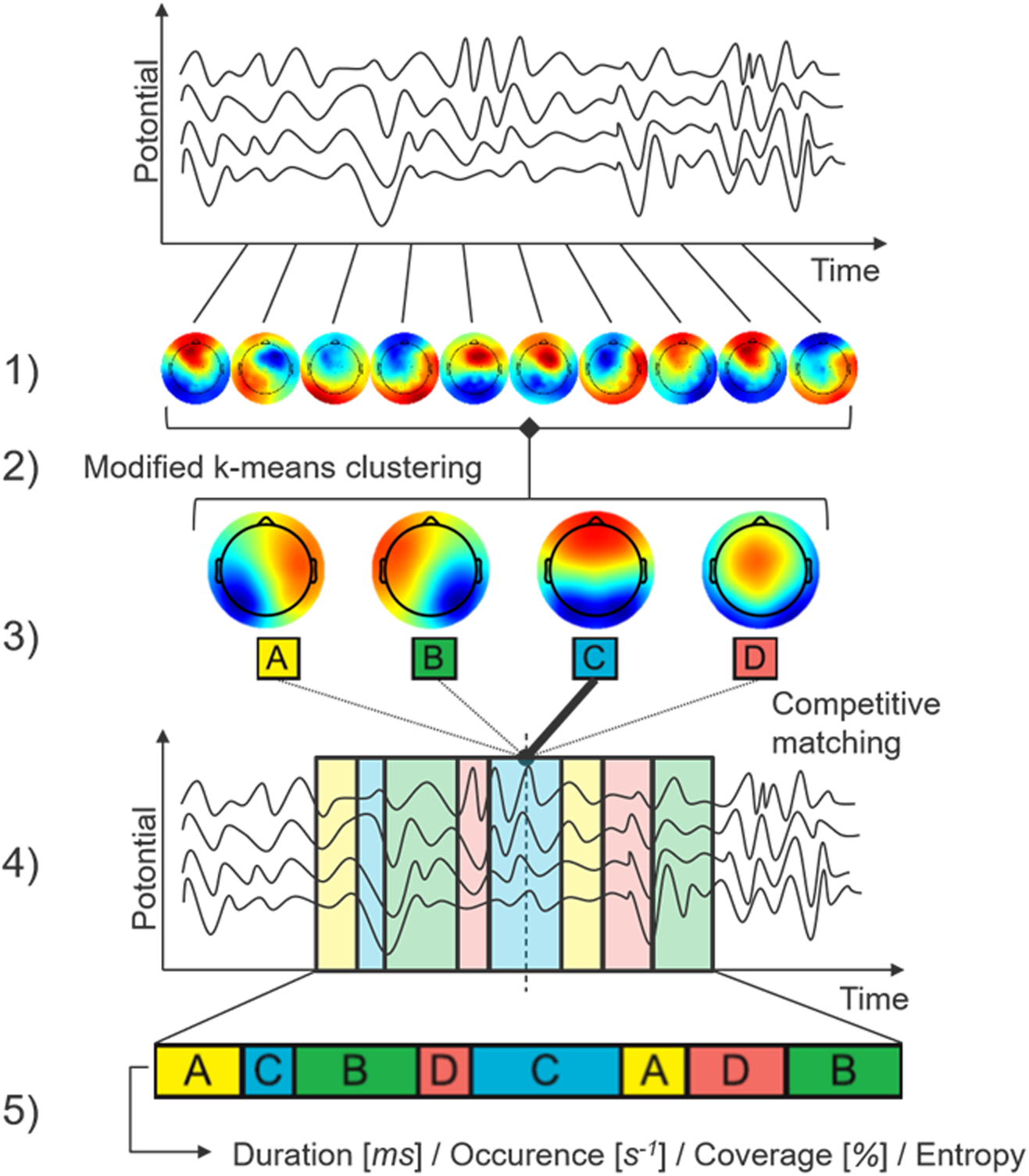
Pipeline for EEG processing.

Microstates were backfitted to the original EEG signal and the resulting sequence of microstates was smoothed using an algorithm^27^ with parameters (window size = 3, smoothing strength = 10) selected to obtain temporal microstate features of HC mimicking those in la Cruz et al.^5^. From the smoothed sequence, temporal microstate features Duration, Occurrence, and Coverage were calculated for each subject. The first and last state of each epoch was ignored, as the true length of that state was unknown. Furthermore, we calculated the transitory feature Sample Entropy of the simulated and permuted sequence. Next, we calculated sample entropy z-scores of microstate sequences standardized with their respective random permutation for different sample lengths. As signal length can influence sample entropy, we accounted for the different numbers of epochs across subjects by estimating the transition and emission matrices of a Hidden-Markov model (HMM) using the Viterbi algorithm^29^. Occurrences of unlabeled states (*K*>4) were reduced to a single “unknown” state *E*. The number of hidden states of the HMM was set equal to the number of states in the sequence, i.e., four or five. Given the estimated transition and emission probabilities, we simulated a sequence of length 20.000. The sequences were then shortened to non-repetitive occurrences only. This way, the sequence *AAAAACCCDDDCCC* resulted in *ACDC*. The shortened sequences were then randomly permuted ten times, such that the same state did not appear twice in direct succession, e.g., *ACDC* can be permuted to *CACD*, but not to *DCCA*. (Supplementary Text S2 for details).

### Statistical Analysis

Between-group differences in demographic variables for FEP, UHR, and HC at baseline were tested using a one-way ANOVA test for continuous variables, and a Chi-squared test of independence or Fisher’s exact test for categorical variables.

For our primary aim, we tested for group differences in temporal and transitional microstate features at baseline using rmANOVA with microstate class as a within-subject factor. Similarly, we tested for group differences in sample entropy z-scores. P-values were adjusted using false discovery rate (FDR).

For our secondary aims, we tested the effect of time on temporal microstate features by performing a LMM with *visit* (three timepoints, baseline, 6 weeks, and 2 years), *microstate class* (A–D) as within-subject factors, and *group* (HC vs. FEP) as a between-subject factor. P-values were adjusted using FDR.

Post hoc two-sample t-tests compared the effect of time *between groups* on the specific microstate class (A, B, C, D) of the temporal features (Duration, Occurrence, Coverage). For testing the effect of time *within group* on the specific microstate class (A, B, C, D) of each the temporal features (Duration, Occurrence, Coverage) we performed a LMM with *visit* (three timepoints, baseline, 6 weeks, and 2 years), and *microstate class* (A–D) as within-subject factors.

As a planned sensitivity test, we examined to what extent medication potentially explained the variance in microstate dynamics of FEP, by correlating antipsychotic medication (recalculated as chlorpromazine equivalent dose, CPZ_Eq) at 6 weeks with microstate A, B, C, and D, and with the *change* in microstate from baseline to 6 weeks.

Finally, we explored associations to symptom levels using a linear model for each microstate class *A* to *D* and stable temporal microstate features (Duration, Occurrence, Coverage), with the three subdimensions of positive, negative, and general symptom scores from PANSS as independent variables, group as a factor, and age and sex as covariates. P-values for model coefficients were calculated using a permutation test and corrected for multiple testing using FDR.

## Results

### Demographic and clinical

Sample demographics at baseline comparing UHR, FEP, and HC are displayed in Table 1. No age or sex differences were observed between FEP, UHR, and HC (*p*=0.71 and *p*=0.83, respectively). Psychotic symptom load of FEP as measured by PANSS total was significantly reduced from baseline to 6 weeks and 2-year follow-up (p<0.001), see clinical details across time in Supplementary Table S1.

**Table 1:** Demographic and clinical data.

Sample demographics at baseline for healthy controls (HC), individuals at ultra-high risk of developing psychosis (UHR), and patients with first episode psychosis (FEP). P-values indicate statistical differences between groups.
| Baseline characteristic | HC<sub>0</sub><br>N = 94 | UHR<sub>0</sub><br>N = 32 | FEP<sub>0</sub><br>N = 47 |  |
| --- | --- | --- | --- | --- |
|  | Mean (SD) |  |  | p-value<sup>1</sup> |
| Age (years) | 23.1 (5.2) | 23.9 (4.5) | 23.1 (5.1) | 0.705 |
| PANSS Total | - | - | 72 (17) |  |
| PANSS Pos |  |  | 18 (4.0) |  |
| PANSS Neg |  |  | 17 (7) |  |
| PANSS Gen |  |  | 37 (9) |  |
| Education (years) | 13.77 (2.25) | 13.09 (2.57) | 12.17 (2.12) | <0.001 |
| SOFAS | - | 43 (7) | 45 (11) | 0.386 |
|  | N (%) |  |  | p-value<sup>1</sup> |
| Gender |  |  |  | 0.830 |
| Female | 44 (47%) | 13 (41%) | 21 (45%) |  |
| Male | 50 (53%) | 19 (59%) | 26 (55%) |  |

<sup>1</sup>One-way ANOVA; Pearson's Chi-squared test

### Methodological results

#### Number of microstates

We found that for microstates A to D, spatial correlation improved significantly until saturation at seven microstates (Supplementary Tables S2 and S3 and Supplementary Figure S1). As a trade-off between a high template spatial correlation and a low number of abundant microstates, we opted for seven microstates (*K*=7) for further analyses.

#### Similarity of microstate topographies

The rmANOVA of spatial correlations with template microstates showed a non-significant group effect (*p*=0.271) and group-microstate interaction (*p*=0.612), indicating similar microstate topographies across groups.

#### Group difference of microstate temporal parameters at baseline comparing HC, UHR, and FEP

The rmANOVA testing for group differences in HC_0_ vs. UHR_0_ vs. FEP_0_ showed insignificant microstate-group interaction for Occurrence, Duration, and Coverage (*p*=0.77, *p*=0.76, and *p*=0.62, respectively), indicating no group differences for any microstate or microstate parameter (Figure 2). On sample entropy, the rmANOVA showed insignificant group effect (*p*=0.587) and group-sample length interaction (*p*=0.936), see Supplementary Table S4 and Supplementary Figure S2.

**Figure 2:**
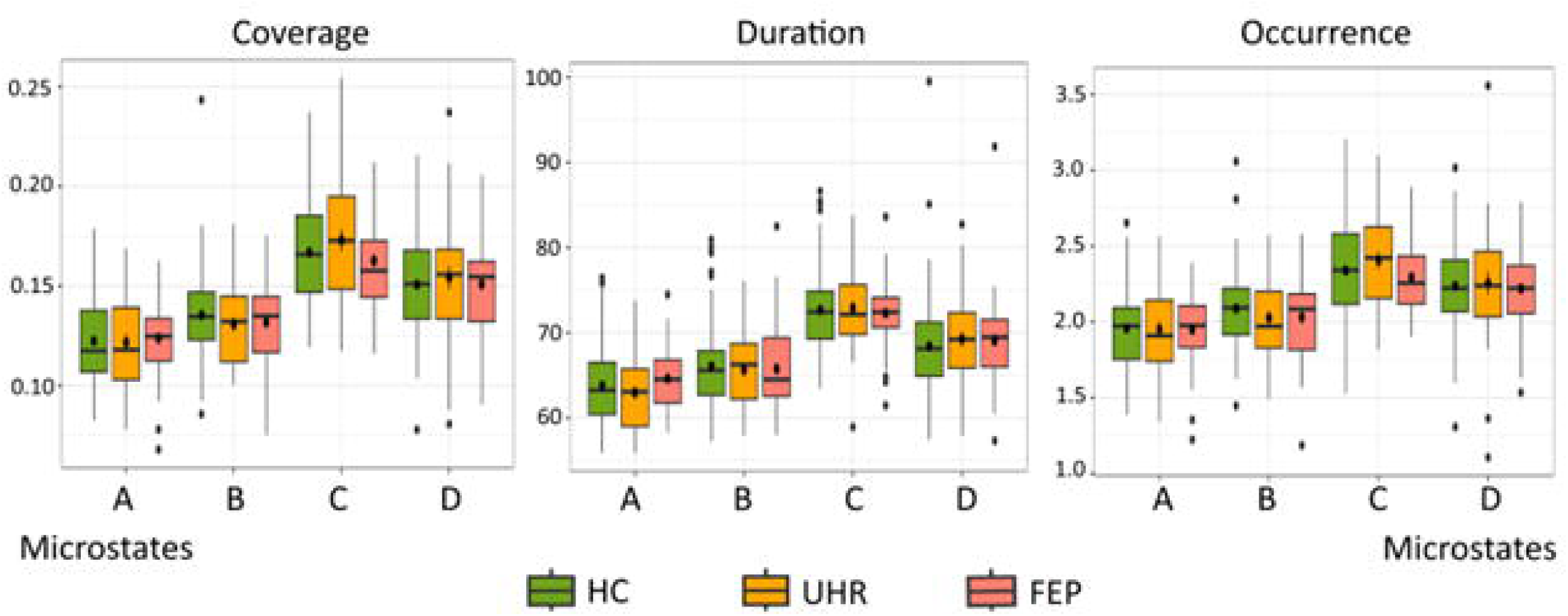
Group difference of Microstate parameters at baseline. *Illustrates the primary analyses of between-group differences of Temporal parameters Coverage, Duration, and Occurrence for each microstate (A-D) at baseline. Abbreviations: FEP: patients with first episode psychosis; HC: healthy controls; UHR: individuals at ultra-high risk for psychosis*.

### Group difference across time

At the 2-year follow-up, we identified a significant effect of time in temporal microstate feature Occurrence (*p*=0.044) and Coverage (*p*=0.036), see Table 2.

**Table 2:** Main LMM Model.

|  | Occurrence |  |  | Duration |  |  | Coverage |  |  |
| --- | --- | --- | --- | --- | --- | --- | --- | --- | --- |
| Term | Estimate | SE | p-value | Estimate | SE | p-value | Estimate | SE | p-value |
| (Intercept) | 1.974 | 0.028 | <0.001* | 63.862 | 0.531 | <0.001* | 0.124 | 0.002 | <0.001* |
| Microstate B | 0.112 | 0.037 | 0.002* | 2.217 | 0.605 | <0.001* | 0.012 | 0.003 | <0.001* |
| Microstate C | 0.354 | 0.037 | <0.001* | 8.429 | 0.605 | <0.001* | 0.042 | 0.003 | <0.001* |
| Microstate D | 0.261 | 0.037 | <0.001* | 4.586 | 0.605 | <0.001* | 0.027 | 0.003 | <0.001* |
| Visit 6W | 0.028 | 0.037 | 0.455 | 0.825 | 0.619 | 0.183 | 0.003 | 0.003 | 0.336 |
| Visit 2Y | 0.013 | 0.043 | 0.765 | 0.741 | 0.721 | 0.304 | 0.002 | 0.004 | 0.609 |
| Group | -0.061 | 0.041 | 0.141 | 0.677 | 0.817 | 0.407 | -0.003 | 0.004 | 0.402 |
| Term | Estimate | SE | p-value | Estimate | SE | p-value | Estimate | SE | p-value |
| Microstate B*Visit 6W | -0.077 | 0.050 | 0.121 | -0.379 | 0.829 | 0.648 | -0.006 | 0.005 | 0.191 |
| Microstate C*Visit6W | -0.033 | 0.050 | 0.514 | 0.083 | 0.829 | 0.920 | -0.001 | 0.005 | 0.745 |
| Microstate D*Visit 6W | -0.022 | 0.050 | 0.659 | -0.273 | 0.829 | 0.742 | -0.001 | 0.005 | 0.805 |
| Microstate B*Visit 2Y | -0.083 | 0.057 | 0.145 | -1.084 | 0.943 | 0.251 | -0.008 | 0.005 | 0.116 |
| Microstate C*Visit 2Y | -0.021 | 0.057 | 0.719 | 0.215 | 0.943 | 0.819 | -0.001 | 0.005 | 0.834 |
| Microstate D*Visit 2Y | -0.028 | 0.057 | 0.619 | 0.158 | 0.943 | 0.867 | -0.002 | 0.005 | 0.760 |
| Microstate B*Group | 0.008 | 0.049 | 0.864 | -0.962 | 0.806 | 0.233 | -0.001 | 0.004 | 0.781 |
| Microstate C:Group | 0.048 | 0.049 | 0.326 | 0.170 | 0.806 | 0.833 | 0.004 | 0.004 | 0.417 |
| Microstate D:Group | 0.048 | 0.049 | 0.323 | -0.026 | 0.806 | 0.974 | 0.003 | 0.004 | 0.455 |
| Visit 6W*Group | 0.016 | 0.042 | 0.695 | -0.085 | 0.749 | 0.910 | 0.001 | 0.004 | 0.774 |
| **Visit 2Y\*Group** | **0.096** | **0.047** | **0.044\*** | 0.016 | 0.864 | 0.986 | **0.008** | **0.004** | **0.036\*** |

\* Significant results, also marked in bold
Abbreviations: 2Y: two years follow-up; 6W: six weeks follow-up; FEP: first episode psychosis; HC: healthy controls; SE: standard error

Sensitivity tests comparing the effect of time between groups on the specific microstate class (A, B, C, D) of the temporal features (Occurrence, Duration, Coverage) revealed a significant group difference between HC and FEP at the 2-years follow-up in Coverage of microstate C (*p*=0.039). No other group differences on microstates were significant, see Supplementary Table S5.

### Within-group effect of time

The post hoc tests showed no effect of time for HC in the temporal features Occurrence, Duration, and Coverage of microstates class A, B, C, and D, respectively (Figure 3). In FEP, there was an effect of time in Occurrence of microstate C between baseline and 2-year (*p*=0.047), on Duration of microstate C between baseline and 2 years (*p*=0.011), as well as Coverage of microstate C between baseline and 2-year (*p*=0.007). No other timepoints or temporal microstate features were significant (for full overview, see Supplementary Table S6).

**Figure 3:**
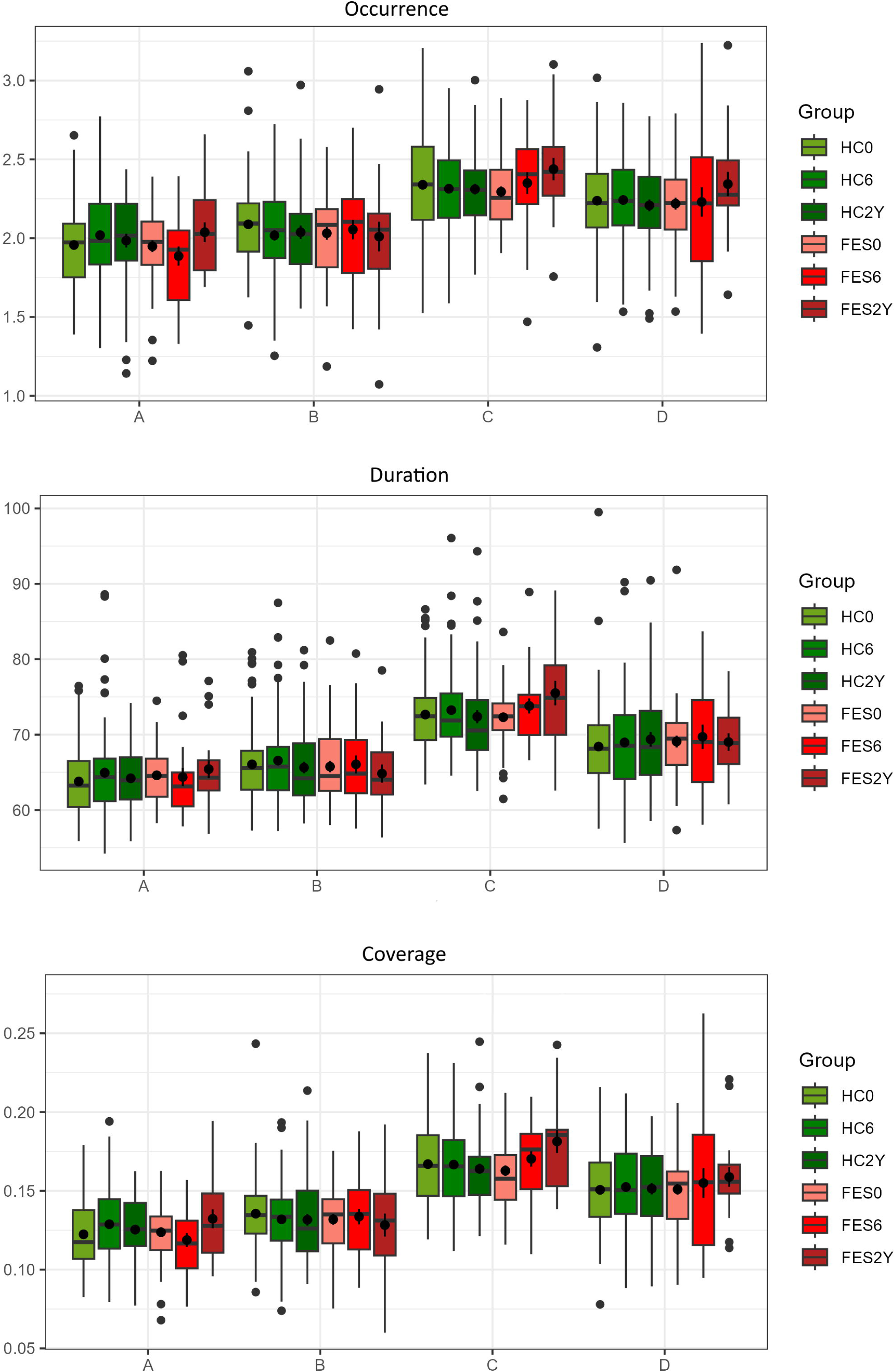
Change in Microstates over time. *illustrates the between- and within-group change of means for temporal microstate features Occurrence (top), Duration (middle) and Coverage (bottom) for each microstate class A, B, C, and D from baseline (HC_0_, FEP_0,_) to post-treatment after 6 weeks (HC_6_, FEP_6_), and to 2 years after baseline (HC_2Y_, FEP_2Y_)*.

### Effect of medication

The correlation test showed no association between CPZ_Eq dose at 6 weeks and microstate Coverage A (*p*=0.765), B (*p*=0.615), C (*p*=0.980), or D (*p*=0.222), all p-values uncorrected. Furthermore, we found no association between CPZ_Eq dose at 6 weeks and change in microstate from baseline to 6 weeks in Coverage A (*p*=0.667), B (*p*=0.243), C (*p*=0.317), or D (*p*=0.290).

### Symptom level correlates

PANSS general symptoms at baseline were negatively associated with Occurrence of microstate D (*r*=-0.016, *p*=0.008). No other PANSS subdimensions were associated with any microstate at any timepoint, see Table 3.

**Table 3:** Associations between microstates and PANSS.

Coefficients ( $r$ ) and significance ( $p$ ), when modelling each microstate (A-D) of temporal features Occurrence, Duration, and Coverage as the dependent variable, with three PANSS subscales (positive, negative, general) as independent variables, and age and gender as covariates.
| Occurrence |  |  |  |  |
| --- | --- | --- | --- | --- |
| PANSS | A | B | C | D |
| Positive | $r=-0.008, p=0.564$ | $r=-0.006, p=0.564$ | $r=-0.009, p=0.564$ | $r=0.015, p=0.564$ |
| Negative | $r=-0.003, p=0.703$ | $r=0.008, p=0.506$ | $r=0.001, p=0.824$ | $r=0.009, p=0.496$ |
| General | $r=-0.001, p=0.980$ | $r=0.005, p=0.710$ | $r=0.001, p=0.808$ | **$r=-0.016, p=0.008$** |
| Duration |  |  |  |  |
| PANSS | A | B | C | D |
| Positive | $r=-0.282, p=0.205$ | $r=-0.040, p=0.922$ | $r=-0.425, p=0.152$ | $r=-0.309, p=0.205$ |
| Negative | $r=-0.138, p=0.468$ | $r=0.025, p=1.000$ | $r=0.015, p=1.000$ | $r=0.012, p=1.000$ |
| General | $r=0.169, p=0.067$ | $r=0.189, p=0.067$ | $r=0.183, p=0.067$ | $r=0.112, p=0.338$ |
| Coverage |  |  |  |  |
| PANSS | A | B | C | D |
| Positive | $r=-0.001, p=0.482$ | $r=-0.001, p=0.623$ | $r=-0.001, p=0.482$ | $r=0.000, p=0.623$ |
| Negative | $r=-0.001, p=0.688$ | $r=0.001, p=0.688$ | $r=0.000, p=1.000$ | $r=0.001, p=0.688$ |
| General | $r=0.000, p=0.623$ | $r=0.001, p=0.430$ | $r=0.000, p=0.465$ | $r=-0.001, p=0.430$ |

Abbreviations: PANSS: Positive And Negative Syndrome Scale

## Discussion

This study aimed to investigate if microstates may be endophenotypic marker for schizophrenia in individuals across the psychosis spectrum^5^ by first comparing HC, UHR, and FEP. Contrary to our expectations, we did not find significant differences in temporal or transitional microstate features between HC, UHR, and FEP at baseline. Overall, we did not reproduce previous cross-sectional observations of increased microstate *C* appearance and decreased microstate *D* appearance in both antipsychotic-naïve^30^ and medicated FEP^5^. However, the average illness duration of patients in the study by da Cruz et al. (2020)^5^ was 12.8 ± 8.2 years, whereas the average duration of illness is less than 1 years at baseline in our FEP sample. The shorter duration of illness may account for the absence of microstate feature alterations, as neurophysiological abnormalities likely become more pronounced with progression to a chronic phase.

Notably, our analyses revealed that contrary to the apparent stability of microstate features in HC over time, we observed an increase of microstate C from baseline to two years after first psychotic episode in FEP, resulting in a significant group difference for coverage of microstate C between HC and FEP at two years follow-up. Although our small sample size and uncorrected post-hoc tests calls for cautious interpretations, our results could indicate that the increased microstate C reported in patients with psychosis is influenced by antipsychotic medication and/or the illness progression.

The FEP patients compared with patients with established schizophrenia in da Cruz et al.^5^ were not antipsychotic-naïve as in our sample. However, we could not confirm a potential effect of antipsychotic medication in the increase of microstate C in FEP over 2 years, as we did not find any association between CPZ_Eq dose and microstates at 6 weeks, as well as change in microstates from baseline to 6 weeks. Importantly, our analyses were limited by the reduced sample size of patients regarding psychopharmacological treatment combined with the EEG examination at the 2-year follow-up, which precluded examination of longitudinal changes.

Sample entropy did not differ between HC, UHR, and FEP. In contrast to Murphy et al^13^ microstate sequence entropy differed between the original and the permuted sequence for small sequence lengths for HC, UHR, and FEP. Differences may be explained by different methodologies, such as the number of microstates or HMM simulated sequences used in this study. Furthermore, the patients in Murphy et al^13^ were almost all medicated, and they reported a clear negative correlation between entropy z-scores and chlorpromazine equivalents, showing that unmedicated patients have z-scores close to zero.

Finally, we explored if microstate temporal parameters were associated with core symptoms as measured by PANSS positive, negative- and general subscales, as reported by eg. Sun et al^31^ by stratifying patient in low- and high symptom levels for comparison with HC. We only observed an association between general symptoms and Occurrence of microstate D, which consistently has been reported as decreased in patients with psychotic disorders^11^. Interestingly, a reduction of occurrence of microstate D has also been reported in patients with general anxiety disorder^32^ and major depressive disorders, along with a negative association between microstate D and level of depression symptoms^33^. Hence, our result may reflect a link between a more general susceptibility for mental illness and reduced microstate D, rather than a biomarker for psychosis as suggested by de Bock et al^6^. In accordance with the review from Wang et al (2022)^4^, the lack of reported associations between microstates and core symptoms across studies calls for cautious interpretation of results, and the need for further research disentangling the longitudinal effects of pharmacological and modulation of EEG microstates by illness progression in large samples are warranted.

### EEG microstate estimation

We identified similar microstate topographies to what is commonly reported in the literature when using seven microstates^8,9^. The first four microstates *A* to *D* were highly correlated with the microstates found by da Cruz et al.^5^. Using only four microstates, we observed that the topography of microstate *D* did not resemble the topography usually found in literature, and in general high inter-subject variability microstates *A*, *B,* and *D* topographies. Spatial correlation with the microstates found by da Cruz et al.^5^ improved significantly for microstates A to D until saturation at seven microstates. Custo & Van De Ville, et al.^34^ use a meta-criterion of 11 optimization criteria to derive the optimal number of seven microstates. Like our findings of varying scalp topography of microstate *D*, they discuss inconsistent labeling of microstate *C* and argue that microstate *C* becomes a combination of two maps, the true map of *C* and a C-like atypical map. Given the topographical similarity of microstates *C* and *D*, it might be that the topographies of these microstates are susceptible to noise. Increasing the number of microstates above four will add atypical microstates, however, might eliminate some of the noise in the typical microstates *A* to *D*. Microstates might be, as suggested by Michel & Koenig^8^, data- and even subject-dependent.

### Limitations

Hyperparameter settings in both preprocessing and microstate analysis challenge direct comparison between studies. Most microstate analyses are performed using CARTOOL^35^, which is closed source. The choice of hyperparameters in the microstate analysis such as smoothing strength and window length can influence temporal results, as temporal smoothing will overpower dominant microstates.

## Conclusion

In conclusion, we found no differences in temporal or transitional microstate features between HC, UHR, or FEP at baseline. Over time, FEP had a significant increase of microstate C after two years, in coverage also compared to HC at two years. Our findings do not reflect that disturbances in EEG microstates constitute endophenotypic markers for psychosis in the early phase of the disease, but rather that illness progression or medication may contribute to the increase. Implications of pharmaceutical modulation of microstates should be further longitudinally investigated. Finally, in this cohort only microstate D were associated with general symptoms, potentially indicating an unspecific disease susceptibility. We furthermore highlight the need for open-source analysis pipelines, such that temporal microstate features of healthy individuals are aligned across studies.

## Supporting information

Ethical permissions

## Data Availability

All data produced in the present study are available with a legal data exchange agreement upon reasonable request to the authors.

## Acknowledgement

We gratefully acknowledge the great effort of our colleagues at the Centre for Neuropsychiatric Schizophrenia Research (CNSR) and Center for Clinical Intervention and Neuropsychiatric Schizophrenia Research (CINS) Mental Health Centre Glostrup and Copenhagen Research Center for Mental Health (CORE), Mental Health Centre Copenhagen. This study was supported by grants from the Lundbeck Foundation (ID: R25-A2701 and ID: R155-2013-16337) and the Dutch Organization for Health Research and Development (ZonMW) Mental Health Program (Grant Agreement No. 60-63600-98-711).

FMM was supported by the Danish Data Science Academy, which is funded by the Novo Nordisk Foundation (NNF21SA0069429) and VILLUM FONDEN (40516)

TDK was supported by a Brain and Behavior Research Foundation 2021 NARSAD Young Investigator Grant (ID 30112). Furthermore, Research Partners Dana and Bill Starling have supported TDK as a Gregory and Tyler Starling Investigator.

## Conflict of interest

Dr. Ebdrup is part of the Advisory Board of Boehringer Ingelheim, Lundbeck Pharma A/S, and Orion Pharma A/S; and has received lecture fees from Boehringer Ingelheim, Otsuka Pharma Scandinavia AB, and Lundbeck Pharma A/S.

Dr. Bojesen received lecture fees from Lundbeck Pharma A/S.

Dr. Glenthøj was the head of the Lundbeck Foundation Centre of Excellence for Clinical Intervention and Neuropsychiatric Schizophrenia Research (CINS) from January 2009 to December 2021, which was partially financed by an independent grant from the Lundbeck Foundation based on international review and partially financed by the Mental Health Services in the Capital Region of Denmark, the University of Copenhagen, and other foundations. All grants are the property of the Mental Health Services in the Capital Region of Denmark and administrated by them. She has no other conflicts to disclose.

## Ethical approval

The studies were approved by the Regional Danish Committee on Health Research Ethics (H-D-2008-088, H-3-2013-149) and conducted in accordance with the ethical standards laid down in the 1964 Declaration of Helsinki and its later amendments.

## Informed consent

All participants provided written informed consent.

## Notes

### Competing Interest Statement

Dr. Ebdrup: I have read the journal's policy and the authors of this manuscript have the following competing interests: I am part of the Advisory Board of Boehringer Ingelheim, Lundbeck Pharma A/S, and Ori-on Pharma A/S and has received lecture fees from Boehringer Ingelheim, Otsuka Pharma Scandina-via AB, and Lundbeck Pharma A/S. Dr. Bojesen: I have read the journal's policy and the authors of this manuscript have the following competing interests: I have received lecture fees from Lundbeck Pharma A/S. Dr. Glenthǿj: I have read the journal's policy and the authors of this manuscript have the following competing interests: I was the head of the Lundbeck Foundation Centre of Excellence for Clinical Interven-tion and Neuropsychiatric Schizophrenia Research (CINS) from January 2009 to December 2021, which was partially financed by an independent grant from the Lundbeck Foundation based on in-ternational review and partially financed by the Mental Health Services in the Capital Region of Denmark, the University of Copenhagen, and other foundations. All grants are the property of the Mental Health Services in the Capital Region of Denmark and administrated by them. She has no other conflicts to disclose. Remainng authors have declared that no competing interests exist.

### Author Declarations

Ethics committee/ Regional Danish Committee on Health Research Ethics (H-D-2008-088, H-3-2013-149) and Regional Ethics Committee of the Capital Region, Denmark (H-D-2009-013) gave ethical approval for this work.

### Summary of Updates

Supplementary files containing ethical permissions updated

