## Supplementary files for "Resting-state electroencephalography microstates in antipsychotic-naïve individuals across the psychosis spectrum"

Text S1

*Microstate analysis*

For each subject, we entered scalp maps at random Global-Field-Power (GFP) peaks into a modified K-means algorithm [30]. GFP is a measure of global brain activity and is calculated by taking the standard deviation of the electrical potential across all electrodes at each time point [31]. High GFP indicates a high signal-to-noise ratio (SNR). The procedure was repeated 6 times to retrieve sets of 4 to 10 microstates (*K* = {4, 5, ..., 10}). Each set of microstates was compared to four template microstates, retrieved from [2], using spatial correlation ($\rho_{s}$) as a similarity measure. Spatial correlation $\rho_{s}$ was calculated using Global Map Dissimilarity (GMD) [31], a strength invariant distance measure of two maps, divided by the number of channels $N_{C}$ (Eq. 1 and 2). We matched microstates using a ‘winner-takes-it-all’ strategy and labeled them accordingly from *A* to *D*. We ignored the polarity of microstate scalp maps during matching. When the number of microstates exceeded the number of template microstates (*K*>4), the remaining microstates remained randomly ordered.

$GMD=\frac{\left\| \frac{x_{n}}{\mathrm{GFP}_{n}}-\frac{x_{n^{'}}}{\mathrm{GFP}_{n^{'}}} \right\|}{N_{C}}$	(1)

$\rho_{s}=1-\frac{\mathrm{GMD}^{2}}{2}$	(2)

After calculating spatial correlation for all microstates in each set and identified microstates *A*, *B*, *C,* and *D*, we estimated the optimal number of microstates by evaluating the measure of fit for each set and by comparing microstates *A* to *D* in each set with the template microstates. For each consecutive set of microstates, we performed an ANOVA using spatial correlations of microstates *A* to *D* with their respective microstates, using microstate class as a factor. The average GEV for each set of microstates was used to evaluate the goodness of fit.

Microstates were backfitted to the original EEG signal by comparing each EEG sample to each microstate using GMD and assigning each sample to its most similar microstate. Global Explained Variance (GEV) is a measure of similarity between each EEG sample *n* and its assigned microstate, calculated as the squared correlation between two maps weighted by the sample fraction of the total GFP. Average GEV is a measure of the goodness of fit for the backfitted microstate sequence to the EEG signal [31]. The resulting sequence of microstates was smoothed using the algorithm by [32] with window size 3 and smoothing strength 10. Smoothing parameters were selected such that temporal microstate features of HC [2]. From the smoothed sequence, temporal microstate feature Duration, Occurrence, and Coverage were calculated for each subject. The first and last state of each epoch was ignored, as the true length of that state was unknown. Note that as the number of microstates exceeds the number of template microstates, only the first four microstates *A* to *D* were considered for statistical analysis. Furthermore, we calculated the transitory feature Sample Entropy of the simulated and permuted sequence. For a given sequence $\left\{ s,s_{i+1}, \ldots,s_{N} \right\}$, we define a subsequence *S* of length *m* where $S_{m}\left( i \right)=\left\{ s,s_{i+1}, \ldots,s_{i+m-1} \right\}$, the tolerance *r*, and the distance measure $d\left[ S_{m}\left( i \right), S_{m}(j) \right]$ for $i\boldsymbol{\neq}j$. Then, Sample Entropy is defined as

$Sample Entropy=-\ln\frac{P}{Q},$	(3)

where *P* is the number of matches $d\left[ S_{m+1}\left( i \right), S_{m+1}(j) \right]<r$ and *Q* is the number of matches $d\left[ S_{m}\left( i \right), S_{m}(j) \right]<r$ [9]. We iterated over $m=\left\{ 1,2,\ldots,8 \right\}$ and chose $r=0.1$. Next, we calculated sample entropy z-scores of microstate sequences standardized with their respective random permutation for different sample lengths.

Signal length can influence sample entropy. To account for different numbers of epochs across subjects, we estimated the transition and emission matrices of a Hidden-Markov model (HMM) using the Viterbi algorithm [33]. Occurrences of unlabeled states (*K*>4) were reduced to a single “unknown” state *E*. The number of hidden states of the HMM was set equal to the number of states in the sequence, i.e., four or five. Given the estimated transition and emission probabilities, we simulated a sequence of length 20,000. The sequences were then shortened to non-repetitive occurrences only. This way, the sequence *AAAAACCCDDDCCC* resulted in *ACDC*. The shortened sequences were then randomly permuted ten times, such that the same state did not appear twice in direct succession, e.g., *ACDC* can be permuted to *CACD*, but not to *DCCA*.

Supplementary Text S2

*Post processing EEG data*

We tested for unbalanced data after preprocessing the EEG signal. We used an ANOVA to test group differences in the number of epochs and interpolated channels.

We estimated the optimal number of microstates by comparing the first four microstates *A* to *D* in each set of microstates with template microstates. For each consecutive set of microstates, we performed an ANOVA using spatial correlations of microstates *A* to *D* with their respective microstates, using microstate class as a factor. The average GEV for each set of microstates was used to evaluate the goodness of fit. Furthermore, to ensure similar microstate scalp topographies across groups, we performed a repeated measure ANOVA (rmANOVA) of spatial correlations of microstates *A* to *D* with their respective template microstates using microstate class as within-subject and group as between-subject factor.

We evaluate robustness of microstate dynamics over time on an individual level and a group level. For assessment of robustness on a group level, we expected stability in microstate dynamics across time points in HC. A rmANOVA with microstates as a between-subject variable and group as a within-subject variable was performed to test for group differences in temporal microstate features among controls (HC_0_ vs. HC_6_ vs. HC_2Y_).

For an assessment of robustness on an individual level, we use corresponding subjects across time-points and performed a group-wise correlation analysis (pairwise, HC_0_ vs. HC_6_ vs. HC_2Y_ as well as FEP_0_ vs. FEP_6_ vs. FEP_2Y_). P-values were estimated using a one-sided permutation test and corrected for multiple comparisons using Bonferroni. Only microstate features showing high temporal robustness were included in the cognitive analysis in order to reduce the number of comparisons.

For all six sets ranging from four to ten microstates, spatial correlation increased when comparing the topographies of microstates A, B, C, and D to their corresponding templates acquired from da Cruz et al. (2020) (Figure ). For consecutive sets of microstates, e.g., sets with four and five microstates, five and six microstates, etc., we tested for differences in spatial correlation between corresponding microstates. We found that for microstates A to D, spatial correlation improved significantly until saturation at seven microstates, Supplementary Table S4 and Supplementary Table S5. As a trade-off between a high template spatial correlation and a low number of abundant microstates, we opted for seven microstates (*K*=7) for further analyses.

Mean spatial correlation with template microstates for microstates *A* to *D* were 0.87, 0.9, 0.96, and 0.88 respectively (*se*<0.02 for all microstates). Trials with one or more microstate correlations below 0.5 were considered outliers (4 out of 355). The average GEV of all seven microstates was 0.75 (se=0.002).

Supplementary Tables

Table S1: Clinical data on patients with psychosis at all timepoints.

| Characteristic | | FEP_0_ N =47 | FEP_6_ N = 24 | FEP_2Y_ N = 19 |  |
| --- | --- | --- | --- | --- | --- |
|  | |  | Mean (SD) | | p-value^1^ |
| PANSS Total | | 72 (17) | 62 (13) | 47 (7) | <0.001 |
| PANSS Positive | | 18 (4) | 14 (4) | 10 (3) |  |
| PANSS Negative | | 17 (7) | 19 (6) | 13 (4) |  |
| PANSS General | | 37 (9) | 29 (6) | 23 (4) |  |
| Chlorpromazine Equivalent (mg/day) | | - | 196 (99) | 253 (170) | 0.272 |
|  | |  | N (%) | | p-value^1^ |
| Antipsychotic  Substance | |  |  |  | 0.011 |
| Amisulpride | | - | 17 (71%) | 3 (15.79%) |  |
| Aripiprazole | | - | 7 (29%) | 1 (5.26%) |  |
| Olanzapine | | - | - | 1 (5.26%) |  |
| Quetiapine | | - | - | 1 (5.26%) |  |
| Risperidone | | - | - | 1 (5.26%) |  |
| Missing | | - | - | 12 (63.16%) |  |

*Table S1 Showing clinical data for patients at all timepoints.*

*^1^One-way ANOVA*

*Abbreviations: FEP: patients with first episode psychosis; PANSS: Positive And Negative Syndrome Scale*

**Table S2 6 vs 7 microstates**

| Microstate | Effect | DFn | DFd | F | p | ges |
| --- | --- | --- | --- | --- | --- | --- |
| A | Nmicro | 1 | 712 | 5.913 | 0.015 | 0.008 |
| B | Nmicro | 1 | 712 | 7.362 | 0.007 | 0.010 |
| C | Nmicro | 1 | 712 | 16.163 | <0.001 | 0.022 |
| D | Nmicro | 1 | 712 | 27.144 | <0.001 | 0.037 |

*Table S2: ANOVA results showing spatial correlation main effects of microstates A to D when comparing 6 and 7 microstates. The spatial correlation increases significantly for all microstates.*

**Table S3 7 vs 8 microstates**

| Microstate | Effect | DFn | DFd | F | p | ges |
| --- | --- | --- | --- | --- | --- | --- |
| A | Nmicro | 1 | 712 | 2.909 | 0.089 | 0.004 |
| B | Nmicro | 1 | 712 | 0.300 | 0.584 | <0.001 |
| C | Nmicro | 1 | 712 | 1.134 | 0.287 | 0.002 |
| D | Nmicro | 1 | 712 | 1.486 | 0.223 | 0.002 |

*Table S3: ANOVA results showing spatial correlation main effects of microstates A to D when comparing 7 and 8 microstates. Spatial correlation does not increase significantly for any microstate.*

**Table S4 ENTROPY**

| **Correlation HC0 – HC6W** | | | | |
| --- | --- | --- | --- | --- |
| **Parameter** | **Length** | **Rho** | **p-value** | **p-adjusted** |
| **Entropy Z** | 3 | 0.21 | 0.031 | 0.155 |
| **Entropy Z** | 4 | -0.02 | 0.552 | 0.783 |
| **Entropy Z** | 5 | -0.09 | 0.783 | 0.783 |
| **Entropy Z** | 6 | -0.07 | 0.727 | 0.783 |
| **Entropy Z** | 7 | -0.06 | 0.707 | 0.783 |
| **Correlation HC0-HC2Y** | | | | |
| **Entropy Z** | 3 | 0.13 | 0.192 | 0.3417 |
| **Entropy Z** | 4 | 0.17 | 0.136 | 0.3417 |
| **Entropy Z** | 5 | 0.12 | 0.205 | 0.3417 |
| **Entropy Z** | 6 | -0.01 | 0.528 | 0.5280 |
| **Entropy Z** | 7 | 0.00 | 0.486 | 0.5280 |
| **Correlation HC6W – HC2Y** | | | | |
| **Entropy Z** | 3 | 0.02 | 0.434 | 0.938 |
| **Entropy Z** | 4 | -0.23 | 0.938 | 0.938 |
| **Entropy Z** | 5 | -0.12 | 0.780 | 0.938 |
| **Entropy Z** | 6 | -0.01 | 0.501 | 0.938 |
| **Entropy Z** | 7 | -0.14 | 0.815 | 0.938 |

| **Correlation FEP0 – FEP6W** | | | | |
| --- | --- | --- | --- | --- |
| **Parameter** | **Length** | **Rho** | **p-value** | **p-adjusted** |
| **Entropy Z** | 3 | -0.08 | 0.631 | 0.8312 |
| **Entropy Z** | 4 | -0.23 | 0.832 | 0.8320 |
| **Entropy Z** | 5 | -0.12 | 0.665 | 0.8312 |
| **Entropy Z** | 6 | 0.09 | 0.366 | 0.8312 |
| **Entropy Z** | 7 | 0.33 | 0.087 | 0.4350 |
| **Correlation FEP0-FEP2Y** | | | | |
| **Entropy Z** | 3 | -0.09 | 0.613 | 0.97 |
| **Entropy Z** | 4 | -0.01 | 0.510 | 0.97 |
| **Entropy Z** | 5 | -0.54 | 0.970 | 0.97 |
| **Entropy Z** | 6 | 0.20 | 0.273 | 0.97 |
| **Entropy Z** | 7 | -0.26 | 0.795 | 0.97 |
| **Correlation FEP6W – FEP2Y** | | | | |
| **Entropy Z** | 3 | -0.40 | 0.812 | 0.975 |
| **Entropy Z** | 4 | 0.45 | 0.157 | 0.785 |
| **Entropy Z** | 5 | -0.15 | 0.609 | 0.975 |
| **Entropy Z** | 6 | -0.44 | 0.830 | 0.975 |
| **Entropy Z** | 7 | -0.64 | 0.975 | 0.975 |

**Table S5 Post- hoc test of group difference in microstates over time comparing HC-FEP**

| **HC6W – FEP6W OCCURRENCE** | | | |
| --- | --- | --- | --- |
| **Microstate** | **T-Statistics** | ***p*-value** | ***p*-adjusted** |
| **A** | 1.944 | 0.060 | 0.529 |
| **B** | -0.525 | 0.602 | 0.938 |
| **C** | -0.479 | 0.635 | 0.841 |
| **D** | 0.125 | 0.902 | 0.938 |
| **HC2Y – FEP2Y OCCURRENCE** | | | |
| **A** | -0.695 | 0.491 | 0.663 |
| **B** | 0.268 | 0.791 | 0.938 |
| **C** | -1.618 | 0.117 | 0.590 |
| **D** | -1.587 | 0.124 | 0.878 |
| **HC6W – FEP6W DURATION** | | | |
| **A** | 0.416 | 0.680 | 0.841 |
| **B** | 0.376 | 0.709 | 0.955 |
| **C** | -0.483 | 0.632 | 0.745 |
| **D** | -0.430 | 0.671 | 0.968 |
| **HC2Y – FEP2Y DURATION** | | | |
| **A** | -0.919 | 0.366 | 0.841 |
| **B** | 0.562 | 0.578 | 0.955 |
| **C** | -1.699 | 0.100 | 0.540 |
| **D** | 0.235 | 0.816 | 0.968 |
| **HC6W – FEP6W COVERAGE** | | | |
| **A** | 1.987 | 0.054 | 0.394 |
| **B** | -0.314 | 0.755 | 0.961 |
| **C** | -0.647 | 0.521 | 0.639 |
| **D** | -0.251 | 0.804 | 0.943 |
| **HC2Y – FEP2Y COVERAGE** | | | |
| **A** | -1.014 | 0.319 | 0.584 |
| **B** | 0.413 | 0.683 | 0.961 |
| **C** | **-2.176** | **0.039** | **0.289** |
| **D** | -1.031 | 0.310 | 0.943 |

**Table S6 Within group effect over time**

| **OCCURRENCE FES0 – FES 6W** | | | | |
| --- | --- | --- | --- | --- |
| **Microstate** | **estimate** | **std.error** | **p.value** | **p_adj** |
| A | -0.060 | 0.067 | 0.370 | 0.850 |
| B | 0.024 | 0.072 | 0.745 | 0.851 |
| C | 0.055 | 0.069 | 0.425 | 0.850 |
| D | -0.029 | 0.071 | 0.688 | 0.851 |
| **OCCURRENCE FES0 – FES 2Y** | | | | |
| A | 0.095 | 0.056 | 0.099 | 0.520 |
| B | -0.011 | 0.084 | 0.897 | 0.936 |
| **C** | **0.132** | **0.065** | **0.047** | **0.520** |
| D | 0.125 | 0.077 | 0.108 | 0.520 |
| **OCCURRENCE FES6W – FES 2Y** | | | | |
| A | 0.151 | 0.089 | 0.096 | 0.520 |
| B | -0.045 | 0.109 | 0.683 | 0.851 |
| C | 0.080 | 0.098 | 0.424 | 0.850 |
| D | 0.114 | 0.125 | 0.368 | 0.850 |

| **DURATION FES0 – FES 6W** | | | | |
| --- | --- | --- | --- | --- |
| **Microstate** | **estimate** | **std.error** | **p.value** | **p_adj** |
| A | -0.223 | 1.088 | 0.839 | 0.875 |
| B | 0.549 | 1.282 | 0.671 | 0.848 |
| **C** | 1.428 | 1.024 | 0.170 | 0.848 |
| D | 0.585 | 1.528 | 0.704 | 0.848 |
| **DURATION FES0 – FES 2Y** | | | | |
| A | 0.992 | 0.938 | 0.300 | 0.848 |
| B | -0.831 | 1.342 | 0.539 | 0.848 |
| **C** | **3.143** | **1.157** | **0.011** | **0.255** |
| D | -0.066 | 1.449 | 0.964 | 0.964 |
| **DURATION FES6W – FES 2Y** | | | | |
| A | 0.824 | 1.609 | 0.612 | 0.848 |
| B | -0.573 | 1.345 | 0.676 | 0.848 |
| C | 0.842 | 1.204 | 0.497 | 0.848 |
| D | -0.692 | 2.088 | 0.742 | 0.848 |

| **COVERAGE FES0 – FES 6W** | | | | |
| --- | --- | --- | --- | --- |
| **Microstate** | **estimate** | **std.error** | **p.value** | **p_adj** |
| A | -0.005 | 0.005 | 0.342 | 0.853 |
| B | 0.002 | 0.006 | 0.765 | 0.893 |
| C | 0.008 | 0.006 | 0.205 | 0.810 |
| D | 0.002 | 0.007 | 0.796 | 0.893 |
| **COVERAGE FES0 – FES 2Y** | | | | |
| A | 0.009 | 0.005 | 0.060 | 0.427 |
| B | -0.003 | 0.007 | 0.664 | 0.893 |
| **C** | **0.017** | **0.006** | **0.007** | **0.173** |
| D | 0.008 | 0.007 | 0.236 | 0.810 |
| **COVERAGE FES6W – FES 2Y** | | | | |
| A | 0.013 | 0.007 | 0.071 | 0.427 |
| B | -0.005 | 0.009 | 0.532 | 0.893 |
| C | 0.011 | 0.008 | 0.201 | 0.810 |
| D | 0.004 | 0.012 | 0.750 | 0.893 |

| **OCCURRENCE HC0 – HC 6W** | | | | |
| --- | --- | --- | --- | --- |
| **Microstate** | **estimate** | **std.error** | **p.value** | **p_adj** |
| A | 0.062 | 0.041 | 0.130 | 0.521 |
| B | -0.065 | 0.037 | 0.080 | 0.520 |
| C | -0.026 | 0.040 | 0.519 | 0.851 |
| D | 0.006 | 0.041 | 0.886 | 0.936 |
| **OCCURRENCE HC0 – HC 2Y** | | | | |
| A | 0.027 | 0.049 | 0.579 | 0.851 |
| B | -0.048 | 0.044 | 0.897 | 0.936 |
| **C** | -0.028 | 0.048 | 0.287 | 0.850 |
| D | -0.021 | 0.041 | 0.618 | 0.851 |
| **OCCURRENCE HC6W –HC 2Y** | | | | |
| A | -0.032 | 0.049 | 0.515 | 0.851 |
| B | 0.020 | 0.053 | 0.712 | 0.851 |
| C | 0.001 | 0.044 | 0.976 | 0.976 |
| D | -0.039 | 0.048 | 0.419 | 0.850 |

| **DURATION HC0 – HC 6W** | | | | |
| --- | --- | --- | --- | --- |
| **Microstate** | **estimate** | **std.error** | **p.value** | **p_adj** |
| A | 1.170 | 0.703 | 0.100 | 0.848 |
| B | 0.539 | 0.649 | 0.409 | 0.848 |
| **C** | 0.542 | 0.629 | 0.391 | 0.848 |
| D | 0.557 | 0.717 | 0.439 | 0.848 |
| **DURATION HC0 – HC 2Y** | | | | |
| A | 0.369 | 0.722 | 0.611 | 0.848 |
| B | -0.251 | 0.735 | 0.734 | 0.848 |
| **C** | -0.168 | 0.826 | 0.839 | 0.875 |
| D | 1.279 | 0.953 | 0.184 | 0.848 |
| **DURATION HC6W – HC 2Y** | | | | |
| A | -0.517 | 0.869 | 0.555 | 0.848 |
| B | -0.819 | 0.893 | 0.362 | 0.848 |
| C | -0.866 | 1.004 | 0.390 | 0.848 |
| D | 0.411 | 1.132 | 0.717 | 0.848 |

| **COVERAGE HC0 – HC 6W** | | | | |
| --- | --- | --- | --- | --- |
| **Microstate** | **estimate** | **std.error** | **p.value** | **p_adj** |
| A | 0.006 | 0.003 | 0.053 | 0.427 |
| B | -0.003 | 0.003 | 0.296 | 0.853 |
| C | -0.000 | 0.004 | 0.921 | 0.959 |
| D | 0.002 | 0.004 | 0.620 | 0.893 |
| **COVERAGE HC0 – HC 2Y** | | | | |
| A | 0.003 | 0.004 | 0.427 | 0.853 |
| B | -0.004 | 0.004 | 0.356 | 0.853 |
| **C** | -0.003 | 0.004 | 0.488 | 0.893 |
| D | 0.002 | 0.004 | 0.679 | 0.893 |
| **COVERAGE HC6W – HC 2Y** | | | | |
| A | -0.003 | 0.004 | 0.418 | 0.853 |
| B | -0.000 | 0.004 | 0.959 | 0.959 |
| C | -0.002 | 0.004 | 0.568 | 0.893 |
| D | -0.001 | 0.005 | 0.819 | 0.893 |

**Supplementary Figures**

**Figure S1**

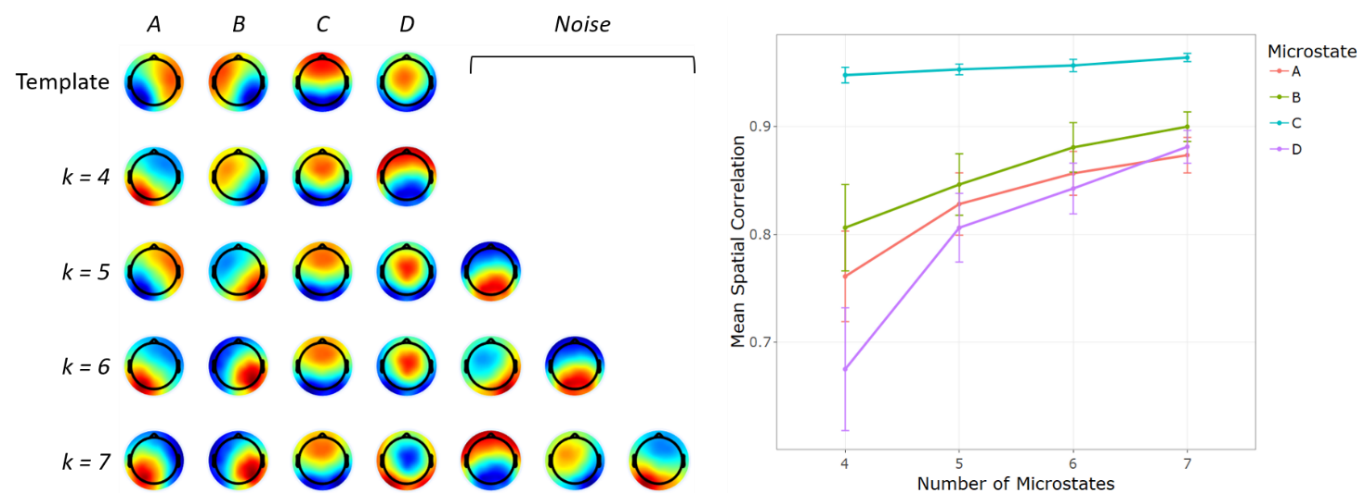

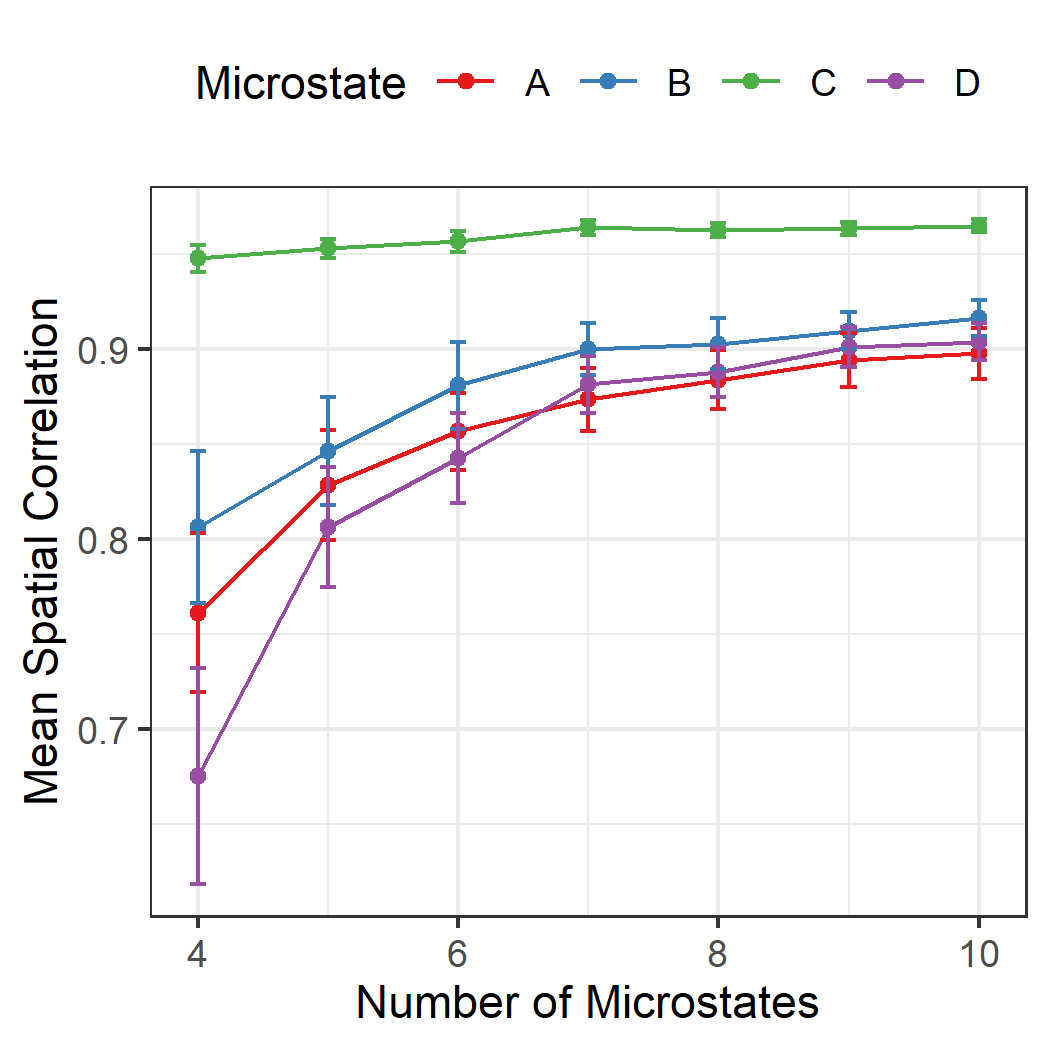

*Figure S1: Left: Mean microstate scalp maps for different number of microstates, sorted according to template microstates. Right: Spatial correlation of microstates A to D with their respective microstate template as a function of the number of microstates obtained during clustering. Microstate C as higher spatial correlation with template microstates than microstates A, B, and D. All microstates improve significantly until saturation at k=7 microstates.*

**Figure S2 Boxplot of entropy values across template length and groups.**

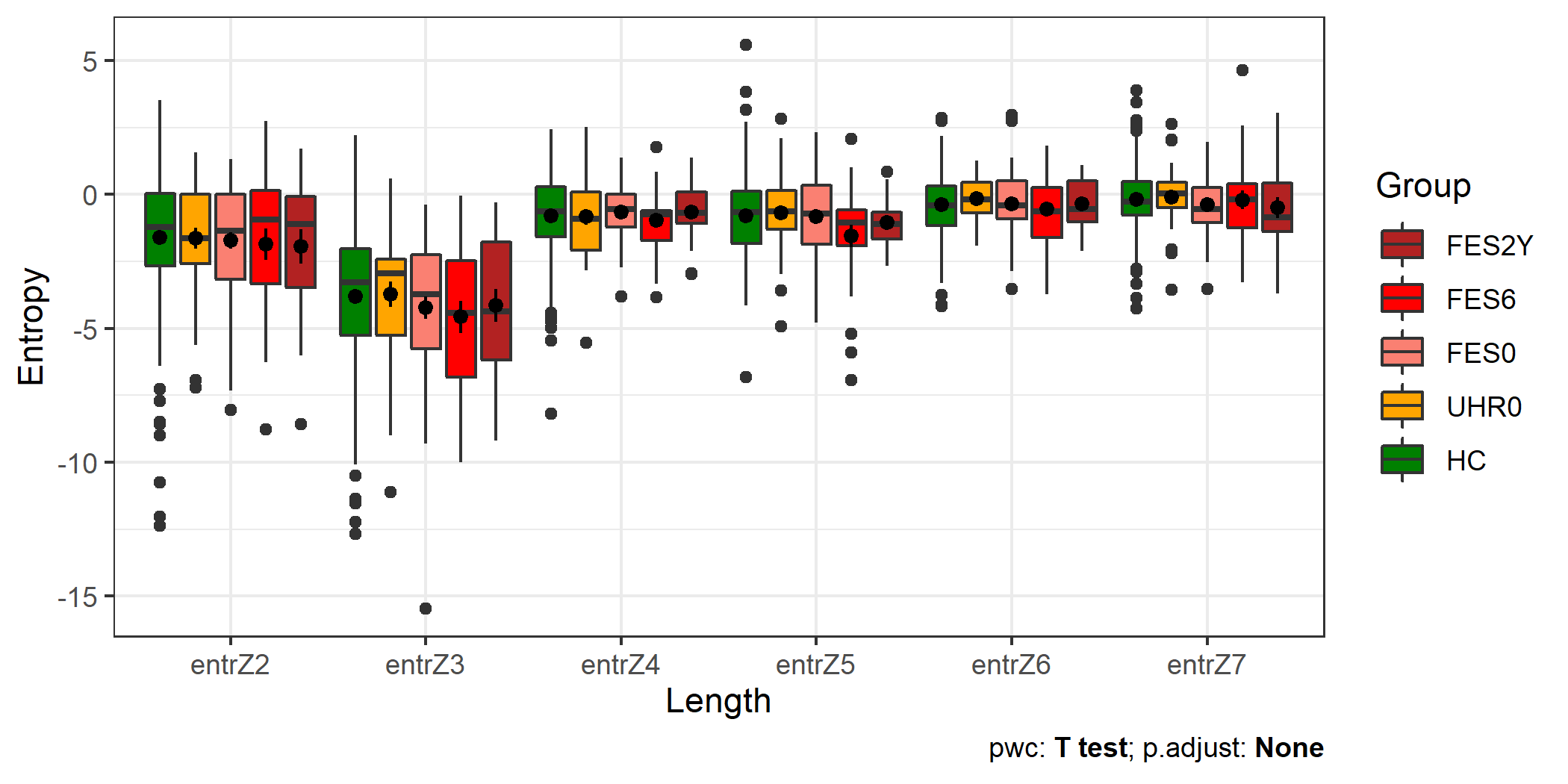

*Supplementary Figure S2: Shows entropy values across template length and groups*
