## Supplementary material for "Resting-state electroencephalography microstates in antipsychotic-naïve individuals across the psychosis spectrum": Ethical permissions: PECANS_ENGLISH.pdf

VEK P1

**Professor, Chief Physician, Doctor of Medicine**
**Birte Glenthøj**
**Psychiatric Center Glostrup**
**Center for Neuropsychiatric Schizophrenia Research**
**Nordre Ringvej 29-67**
**2600 Glostrup**

**Potential biomarkers predicting treatment response and outcome in schizophrenia**
**Final approval**
The decision has been made pursuant to Act No. 593 of June 14, 2011 on the ethical treatment of health science research projects, as subsequently amended.

I confirm receipt of the email dated December 23, 2015, in response to the decision of December 15, 2015, which set conditions for the approval of the project.

The conditions for approval are considered fulfilled. The project is therefore finally approved.

The approval is valid until June 1, 2018 and covers the following documents:

- Study protocol, version 2, 23/12/2015
- Guidelines for oral participant information, version 1, 03/11/2015
- Participant information, version 2, 23/12/2015
- Informed consent form, version 1, 03/11/2015
- Cover letter to participants, version 1, 03/11/2015
- Questionnaires approved for use in the study:
    - SSP-UKU
    - Quality of Life
    - SWM-s
    - EHI

The approval applies to the registered trial sites and the registered principal investigator in Denmark.

Initiating the project in violation of the approval may result in fines or imprisonment, cf. Section 41 of the Committee Act.

### Changes

If significant changes are made to the protocol materials during the course of the project, these must be reported to the committee in the form of amendment protocols. Changes may only be implemented after approval by the committee, cf. Section 27(1) of the Committee Act.

Notification of amendment protocols must be submitted electronically at www.drvk.dk using the assigned registration number and access code.

Significant changes include those that may affect participant safety, interpretation of the scientific documentation underlying the project, or the conduct or management of the project. Examples include changes in inclusion/exclusion criteria, study design, number of participants, study procedures, treatment duration, efficacy parameters, changes regarding investigators or trial sites, and substantive changes to written participant information.

If new information leads the researcher to consider changing procedures or stopping the trial, the committee must be informed.

---

### Adverse reactions and events

#### Ongoing reporting

The committee must be notified immediately if suspected serious, unexpected adverse reactions or serious events occur during the project, cf. Section 30(1) of the Committee Act. The report must include comments on any consequences for the trial. Only adverse reactions and events occurring in Denmark must be reported. Notification must occur no later than 7 days after the sponsor or principal investigator becomes aware of the case.

A reporting form is available at www.dnvk.dk. The form and attachments can be submitted electronically using a digital signature or on CD-ROM.

#### Annual reporting

Once a year throughout the trial period, the committee must receive a list of all suspected serious (expected and unexpected) adverse reactions and serious events that occurred during the trial, along with a report on participant safety, cf. Section 30(2) of the Committee Act.

The material must be in Danish or English.

The reporting form is available at www.dnvk.dk. The form and attachments can be submitted electronically using a digital signature or on CD-ROM.

---

### Completion

The principal investigator and any sponsor must notify the committee no later than 90 days after the project ends, cf. Section 31(1) of the Committee Act. The project is considered completed when the last participant has finished.

If the project is terminated earlier than planned, a justification must be sent to the committee no later than 15 days after the decision, cf. Section 31(2) of the Committee Act.

If the project does not commence, this and the reason must be reported to the committee.

The committee requests a copy of the final research report or publication, cf. Section 28(2) of the Committee Act. We emphasize that there is an obligation to publish both negative, positive, and inconclusive trial results, cf. Section 20(1)(8) of the Committee Act.

The obligation to report the final trial and report rests jointly with the principal investigator and any sponsor.

---

**Supervision**

The committee monitors that the project is conducted in accordance with the approval, cf. Sections 28 and 29 of the Committee Act.

---

**Signature on the consent form**

The committee notes that the principal investigator may delegate the duty to sign the consent form to the person conducting the oral information session. In such cases, there must be a written delegation at the trial site.
