## Supplementary material for "Resting-state electroencephalography microstates in antipsychotic-naïve individuals across the psychosis spectrum": Ethical permissions: PECANSII_ENGLISH.pdf

VEK P2

**Name:**
Professor, Dr. Med. Birte Glenthøj

Center for Neuropsychiatric Schizophrenia Research (CNSR),
Psychiatric Center Glostrup

Ndr. Ringvej 29-67, 2600 Glostrup


**Sent to:**
Kirsten Borup Bojesen

**Approval Authority:**
Region Hovedstaden, IMT
Borgervænget 7
2100 Copenhagen Ø

**Data Controller:**
Region Hovedstaden
Kongens Vænge 2
3400 Hillerød

---

**Approval**

Updated with new data processor
Regarding notification of:
**"(PECANS II) Pan European Collaboration on Antipsychotic Naïve Schizophrenia II (PECANS II) study"**

The above-mentioned project was reported on 18-12-2013 under the umbrella notification scheme between Region Hovedstaden and the Danish Data Protection Agency.
Approved by Lilian Schelde Baunbæk, Information Security, IMT.

The notification states that you are responsible for the project's data. Data processing is scheduled to begin on 01-01-2014 and is expected to end on 31-12-2033.

Your notification of data processing titled **"(PECANS II) Pan European Collaboration on Antipsychotic Naïve Schizophrenia II (PECANS II) study"** is now approved and registered under Region Hovedstaden's umbrella notification for health science research with journal no.: **2007-58-0015**.

Your local journal number under Region Hovedstaden's umbrella notification is: **RHP-2013-026**, with I-Suite no: **02579**, and this must be used for inquiries, changes, or applications for project extension.

The project will include examination of biological material.

The project will receive information from another approved research project in Region Hovedstaden, with journal no.: **PSV-2001-04 Danish Psychiatric Biobank / I-Suite 00422 – ID no 2001-54-0798**.

---

### Data Processors

Data will be processed by the following data processors:

- Laboratory Filadelfia v/Jan Borg Rasmussen
- Clinical Biochemistry Department, Aarhus, by Project Administrator Uffe Lund Lystbæk
- Statens Serum Institut (SSI), by Gunnar Houen, Professor, Dr. Scient
- Faculty of Science, University of Copenhagen, by Jens Lykkesfeldt, Professor

The project is approved to receive data through:
Access to relevant systems via approval from: **Research Ethics Committee**

---

### Permission

You are hereby granted permission to conduct the project under the following conditions:

- The permission is valid until: **31-12-2033**
- If you have not extended the permission by this date, Region Hovedstaden assumes that the project is completed and that the data has been deleted, anonymized, destroyed, or transferred to an archive.
- Please note that any data processing, including passive storage, of personal data after the permission expires is a violation of this permission.

---

### Changes to the Project

Significant changes, such as if your project:

- is not completed as planned

- is terminated early
- adds external data processors
- adds other personally identifiable data

must be reported to the local contact person for data notifications. Remember to provide your local journal number and I-Suite number as stated at the top of this email.

---

**Electronic Data – Security Requirements**

- Identification data must be encrypted or replaced with a code number or similar. Alternatively, all data can be stored encrypted. Encryption keys must be stored securely and separately from personal data.
- Note that encrypted or coded data is only **pseudonymized**. Data is only fully anonymized when it is technically and humanly impossible to identify the actual person.
- Access to project data must only occur using a confidential password.
- When transferring personally identifiable data via the internet or other external networks, appropriate security measures must be taken to prevent unauthorized access. Data must be encrypted throughout transmission. For internal networks, ensure that unauthorized persons cannot access the data.
- Data must not be stored on a laptop. Instead, obtain an encrypted USB key from IMT for storing your data.
- Databases must not be stored on a personal drive, typically an H:\ drive.
- Removable storage media, data backups, etc., must be stored securely and locked to prevent unauthorized access.

Further reference is made to the rules in the **Security Executive Order BEK. no. 528**, which all public authorities must comply with.
Link to the Security Executive Order: https://www.retsinformation.dk/Forms/R0710.aspx?id=1002
