## Supplementary material for "Resting-state electroencephalography microstates in antipsychotic-naïve individuals across the psychosis spectrum": Ethical permissions: UHR_ENGLISH.pdf

**Permission**
Regarding notification of:
"NEURAPRO-E – North America, Europe, Australia Prodromal Study. A multicenter randomized trial of omega-3 fatty acids and cognitive behavioral case management for symptomatic patients at special risk of early progression."

The above-mentioned project was reported on March 20, 2018, under the umbrella notification scheme between the Capital Region of Denmark and the Danish Data Protection Agency.

Permission has been granted by Heidi Dorthe Jensen, Knowledge Center for Data Notifications, Legal Unit, Rigshospitalet.

The notification states that you are responsible for the project's data. Processing of the information is intended to begin on March 20, 2018, and is expected to end on February 24, 2022.

Your notification of data processing titled "NEURAPRO-E – North America, Europe, Australia Prodromal Study. A multicenter randomized trial of omega-3 fatty acids and cognitive behavioral case management for symptomatic patients at special risk of early progression" is now approved and registered under the Capital Region's umbrella notification for health science research with ref. no.: 2012-58-0004.

Your local ref. no. under the Capital Region's umbrella notification is: VD-2018-54, with I-Suite no.: 6263, and it must be used for inquiries, changes, or applications for extension of your project.

The project will include examination of biological material. Analyses have been conducted.
Version 1.0 2017

Legal basis for data is provided through: Consent form and Scientific Ethics Committee.

This permission is solely a permission to process personal data in connection with the implementation of the project. The permission does not entail an obligation for authorities, companies, etc., to disclose any information to you for use in the project.

The project was reported as private research in 2009 under ref. no. 2009-41-3128 and at the latest extension with new ref. no. 2014-41-2861. The project's permission expired on August 1, 2015, and at that time the researcher was in contact with the Data Protection Agency, which stated that an extension was not necessary since the project was reported as private research and approved by the Ethics Committee. It is an error that they were not made aware that it should have been registered under the Region's umbrella notification.

The data controller contacted the Data Protection Agency on January 4, 2018, as data was still being processed, to request permission to continue the project under the Capital Region's umbrella permission, even though the original permission had expired. The data controller states that they acted in good faith. In an email dated January 17, 2018, the Data Protection Agency recommended that [data controller] instead report the project to the Capital Region. It also appeared from the correspondence between the researcher and the Data Protection Agency that the project had apparently continued without notification for a period after the notification with the Data Protection Agency had expired. It cannot be documented how data was stored during that period.

The Legal Unit assesses that data from "NEURAPRO-E – North America, Europe, Australia Prodromal Study. A multicenter randomized trial of omega-3 fatty acids and cognitive behavioral case management for symptomatic patients at special risk of early progression" are valuable data that would be difficult and time-consuming to collect again. The data are of societal and health significance, which also contributes to this assessment.

It has been agreed that the Data Protection Agency's reference numbers are noted in the Region's register, and previous permissions from the Data Protection Agency are attached along with a telephone note about the notification.

**Permission**
You are hereby granted permission to store data under the following conditions:

The permission is valid until February 24, 2018.
If you have not obtained an extension of the permission by this date, the Capital Region assumes that the project is completed and that the information has either been deleted, anonymized, destroyed, or transferred to an archive.

Please note that any data processing, including passive storage, of personal data after the permission expires is a violation of this permission.

The permission applies exclusively to the disclosure of data to the above-mentioned approved project. If data from the above-mentioned project is to be disclosed to another project with a different purpose, separate permission must be obtained. Please note that according to Section 46 of the Health Act, it is not permitted for a person associated with the project to independently collect data from source systems, such as PAS systems. This also applies to internal and external collaborators and monitors.

The permission is furthermore granted on the condition that the researcher obtains relevant permissions. Upon request from the Capital Region, documentation of the permission must be presented.

Version 1.0 2017

---

**Changes to the Project**

Significant changes, such as if your project:

- is not completed within the planned timeframe

- is completed earlier than planned
- involves external data processors
- adds other personally identifiable data

must be reported to the local contact person for data notifications. Remember to provide your local reference number and I-Suite number, as stated at the top of the email.

---

**Electronic Data – Security Requirements**

Identification data must be encrypted or replaced with a code number or similar. Alternatively, all data can be stored in encrypted form. The encryption key, code key, etc., must be stored securely and separately from the personal data.

Please note that coded data is only partially anonymized.
Data is only considered anonymized when it is no longer possible to use it to identify specific individuals—whether by human, technical, or other means.
Encrypted data is still considered identifiable data.

Access to project data may only occur using a confidential password.
In accordance with the Security Executive Order BEK 528 of 15/06/2000, there will be automated logging of users and the time of data processing.

When transferring personally identifiable information via the internet or other external networks, appropriate security measures must be taken to prevent unauthorized access. The information must, at a minimum, be encrypted throughout the entire transmission. When using internal networks, it must be ensured that unauthorized persons cannot access the information.

The information must not be stored on a laptop computer.
For temporary data storage, the project can obtain an encrypted USB key from CIMT, where you can keep your information.
Version 1.0 2017

Data must only be stored in the folders specified and approved in the security schema on the Capital Region's network.
Removable storage media, data backups, etc., must be stored securely under lock and key so that unauthorized persons cannot access the information.

Reference is also made to the rules in the Security Executive Order BEK no. 528 of 15/06/2000, which all public authorities must comply with.
Link to the Security Executive Order: https://www.retsinformation.dk/Forms/R0710.aspx?id=1002

---

### Duty to Inform the Data Subject

If information is to be collected from the data subject (through interview, questionnaire, clinical or paraclinical examination, treatment, observation, etc.), detailed information about the project must be provided/sent. The data subject must be informed of the name of the data controller, the purpose of the project, that participation is voluntary, and that consent to participate can be withdrawn at any time.

If the information is to be disclosed for use in another scientific or statistical context, the purpose of the disclosure and the recipient's identity must also be stated.

The data subject must also be informed that the project has been reported to the Danish Data Protection Agency under the Personal Data Act, via the Capital Region as a delegated authority, and that the Data Protection Agency has set specific conditions for the project to protect the privacy of the data subject.
